# Differential Effects of Race/Ethnicity and Social Vulnerability on COVID-19 Positivity, Hospitalization, and Death in the San Francisco Bay Area

**DOI:** 10.1101/2022.01.04.22268760

**Authors:** Wendy K. Tam Cho, David G. Hwang

## Abstract

**BACKGROUND:** Higher COVID-19 incidence and morbidity have been documented for US Black and Hispanic populations but not as clearly for other racial and ethnic groups. Efforts to elucidate the mechanisms underlying racial health disparities can be confounded by the relationship between race/ethnicity and socioeconomic status.

**OBJECTIVE:** Examine race/ethnicity and social vulnerability effects on COVID-19 outcomes in the San Francisco Bay Area, an ethnically and socioeconomically diverse region, using geocoded patient records from 2020 in the University of California, San Francisco Health system.

**KEY RESULTS:** Higher social vulnerability, but not race/ethnicity, was associated with less frequent testing yet a higher likelihood of testing positive. Asian hospitalization rates (11.5%) were double that of White patients (5.4%) and exceeded the rates for Black (9.3%) and Hispanic patients (6.9%). A modest relationship between higher hospitalization rates and increasing social vulnerability was evident only for White patients. Hispanic patients had the highest years of expected life lost due to COVID-19.

**CONCLUSIONS:** COVID-19 outcomes were not consistently explained by greater social vulnerability. Asian individuals showed disproportionately high rates of hospitalization regardless of social vulnerability status. Study of the San Francisco Bay Area population not only provides valuable insights into the differential contributions of race/ethnicity and social determinants of health to COVID-19 outcomes but also emphasizes that all racial groups have experienced the toll of the pandemic, albeit in different ways and to varying degrees.

## Introduction

Previous national [1, 2], regional [3], and hospital-based [4] studies in the United States (US) have demonstrated a pattern across a variety of states and localities of Black and Hispanic individuals testing positive for COVID-19 at higher rates than White individuals. Given the enormous geographic and contextual diversity across the US, each of these studies provides some insight into the complex relationship between race and the COVID-19 pandemic as well as the larger phenomenon of general racial health disparities.

Conducting racial health disparity studies across different geographic and demographic settings is critical for at least two reasons. First, since each community is unique in composition and history, it is important to understand how contextual differences may affect the manifestation of racial health disparities and their associated underlying contributors. Second, in many US communities race and socioeconomic status (SES) are closely tied, making it difficult to separate their relative contributions to health disparities. Accordingly, US communities characterized by both high racial and ethnic diversity as well as high socioeconomic diversity are of particular interest for examining the differential contributions of race and ethnicity and socioeconomic factors to health disparities. As a novel, widespread, and contemporaneous disease entity with discrete onset, the COVID-19 pandemic provides a unique and invaluable opportunity to study racial inequities in health care delivery and outcomes in the US [5].

The San Francisco Bay Area is the 12th largest metropolitan statistical area in the US with approximately 7.7 million inhabitants, according to the 2020 US Census. The geographic region is one of the most racially and ethnically diversion metropolitan areas in the country [6], and its residents exhibit multifaceted diversity. The Bay Area is majority non-White with 36% Whites, 6% Blacks, 28% Asians, 24% Hispanics, and 6% other (2020 US Census). In addition to exhibiting high racial and ethnic diversity, the population in this geographic area also demonstrates marked income inequality, as evidenced by an 11-fold income difference between households in the 90th and the 10th income percentiles. Moreover, fully one-third of the households are characterized as being very low income [7, 8].

The composition of the very low income group in the San Francisco Bay Area is also racially and ethnically diverse (35% Hispanic, 25% Asian, 26% White, and 10% Black). In the Bay Area, the majority of Black (50%) and Hispanic (54%) residents are in the very low income group and a minority are in the high income group (18% of Blacks and 15% of Hispanic residents). In contrast, Asian residents are more evenly distributed across the income spectrum, with 31% of the group occupying the very low income group and 36% in the high income group [8]. These differences between the racial and ethnic groups reflect, in part, each group’s unique history (e.g., intergenerational trauma from slavery and lynching, crowded housing and poor industry working conditions, immigration exclusions, and refugee policies) [9, 10, 11]. Many inequities have targeted one particular group. Indeed, the inequities and resulting socioeconomic heterogeneity are unevenly shared. Nonetheless, while the historical, contextual, and etiologic underpinnings of health inequalities may differ, the evolution and current characteristics each of these groups share some common themes that contribute to present-day health disparities experienced by many individuals of minority groups.

To further our understanding of why racial health disparities exist and how to ameliorate them, we investigated the demographics of COVID-19 positivity, hospitalization, and death in the San Francisco Bay Area. In particular, the relative decoupling of race and ethnicity and SES among Bay Area Asians presents a unique opportunity to help unravel the complex and typically confounding interrelationship between race and ethnicity and other social determinants of health on observed outcomes for COVID-19.

## Methods

Under an Institutional Review Board (IRB) approved protocol (IRB #20–30545), an analysis was performed of all the electronic health records (EHR) of patients within the University of California San Francisco (UCSF) Health system, a large multi-hospital, multi-clinic academic health care system in the San Francisco Bay Area. Except for address data, all other identifying data were redacted for confidentiality purposes.

Inclusion criteria were threefold. First, the patient’s residence as listed in the EHR is in one of the nine Bay Area counties: Alameda, Contra Costa, Marin, Napa, San Francisco, San Mateo, Santa Clara, Solano, or Sonoma. A benefit of this restriction is that it reduces referral bias effects, in which patients referred from outside UCSF’s immediate Bay Area catchment area for tertiary care tend to skew towards having poorer health as well as being more indigent. Second, the patient self-reported his or her racial and ethnic identification as non-Hispanic Black (hereafter, Black), non-Hispanic White (hereafter, White), non-Hispanic Asian (hereafter Asian), or Hispanic or Latino (hereafter Hispanic). The small number of patients (approximately 1%) self-reporting as Native Hawaiian or Other Pacific Islander were included with the Asian group. We excluded records from individuals who self-reported as multi-racial (2.6%), in which no race or ethnicity was reported (8.4%), or in which race was reported as American Indian or Alaska Native (less than 1%). While these groups are of potential interest, their small numbers in our data set precluded drawing meaningful conclusions from the additional analysis performed for the other, larger racial and ethnic groups. Significant selection bias due to exclusion of this small proportion of records is possible, but likely to be low. Third, the patient had to have undergone reverse transcriptase polymerase chain reaction (RT-PCR) testing for SARS-CoV-2 at least once in calendar year 2020. This time period was chosen to exclude the influence of vaccinations, first doses of which were available to less than 1% of the study population as early as mid-December 2020 and second doses of which would not have been completed until January 2021 for the earliest vaccinated individuals. Since RT-PCR testing was not widely available during the first several months of 2020, relatively few tests were recorded in January 2020 and February 2020, but the number increased steadily starting in March 2020. The earliest test result in our data set is from January 4, 2020, and the latest test result is from December 31, 2020.

To gain a sense of socioeconomic status absent the ability to capture individual-specific measures not recorded in the EHR (such as household income), we utilized the Centers for Disease Control (CDC) Social Vulnerability Index (SVI) recorded for the census tract corresponding to the patient’s place of residence. The CDC SVI incorporates US Census data to determine the social vulnerability of every census tract in the US. We mapped this measure to each patient record based on patient address listed in the EHR. The SVI summarizes a broad set of data from four different types of measures: socioeconomic factors, household composition and disability factors, minority status and language factors, and housing type and transportation factors [12]; the higher the SVI value, the more vulnerable are the individuals within the census tract. In our data, CDC SVI was highly correlated (*ρ* = 0.87) with measures of poverty. Thus, while not a direct measure of SES per se, SVI is a reasonable proxy for direct measures of SES such as household income. Higher CDC SVI mapped to the county level has been reported to correlate with higher incidence and mortality rates of COVID-19 in the United States [13].

## Results

### Racial and Ethnic Demographics

The racial and ethnic composition of the study population, the US population, and the San Francisco Bay Area population is shown in Table 1. The demographics of the Bay Area differ from the general US population in having greater racial diversity, with far fewer White individuals, many more Asian persons, lower proportions of Black individuals, and higher proportions of Hispanic persons. The study population has a racial and ethnic distribution that differs from that of both the US population and the Bay Area population. The proportion of White individuals in the study population (57.9%) is lower than that in the US (76.3%) but notably higher than that in the Bay Area (35.9%). Asian individuals, by contrast, constitute a much higher proportion of the study population (15.4%) than of the US (6.1%) but are significantly underrepresented relative to their proportion in the Bay Area (28.3%).

**Table 1:** Racial/Ethnic Distribution of the Study Population in Comparison to the US and San Francisco Bay Area.

|  | White | Black | Hispanic | Asian |
| --- | --- | --- | --- | --- |
| US population | 76.3% | 13.4% | 18.5% | 6.1% |
| San Francisco Bay Area population | 35.9% | 5.6% | 24.4% | 28.3% |
| Study Population | 57.9% | 7.4% | 19.0% | 15.4% |

### COVID-19 Testing Frequency

Table 2 shows the percentage of total tests and the percentage of positive tests ascribed to each race and ethnicity. No significant association between race and the frequency of COVID-19 testing was observed. The distribution of race and ethnicity among the COVID-19 testing group (Table 2) largely mirrored the racial proportions in the overall UCSF patient pool (Table 1). White and Hispanic individuals underwent testing at slightly lower rates than their overall UCSF patient pool proportions, while Black and Asian individuals underwent testing at somewhat higher rates. These differences were small. For all racial and ethnic groups, the absolute deviations from the UCSF patient pool percentages were no more than 3%.

**Table 2:** COVID-19 Testing and Positivity Rates by Race/Ethnicity.

|  | White |  | Black |  | Hispanic |  | Asian |  |
| --- | --- | --- | --- | --- | --- | --- | --- | --- |
| Testing | 55.69% | (73,503) | 8.19% | (10,815) | 17.54% | (23,155) | 18.34% | (24,205) |
| Positivity Rate | 2.54% | (1,866) | 5.38% | (582) | 11.76% | (2,722) | 3.68% | (890) |

Absolute number shown in parentheses.

By contrast, an inverse relationship was observed between the CDC SVI and the frequency of undergoing COVID-19 testing. Figure 1 depicts the percentage of the total UCSF population (solid line) occupying each CDC SVI decile (SV1 = least vulnerable; SV10 = most vulnerable), as well as the SVI distribution of individuals undergoing COVID-19 testing (dashed line). In the decile representing the least socially vulnerable, individuals underwent testing at higher rates than their corresponding population proportion. For other SVI deciles, individuals with lower vulnerability tended to tested at slightly higher rates than their population proportion, whereas those in the more vulnerable deciles tested at moderately lower rates than their population proportion.

**Figure 1:**
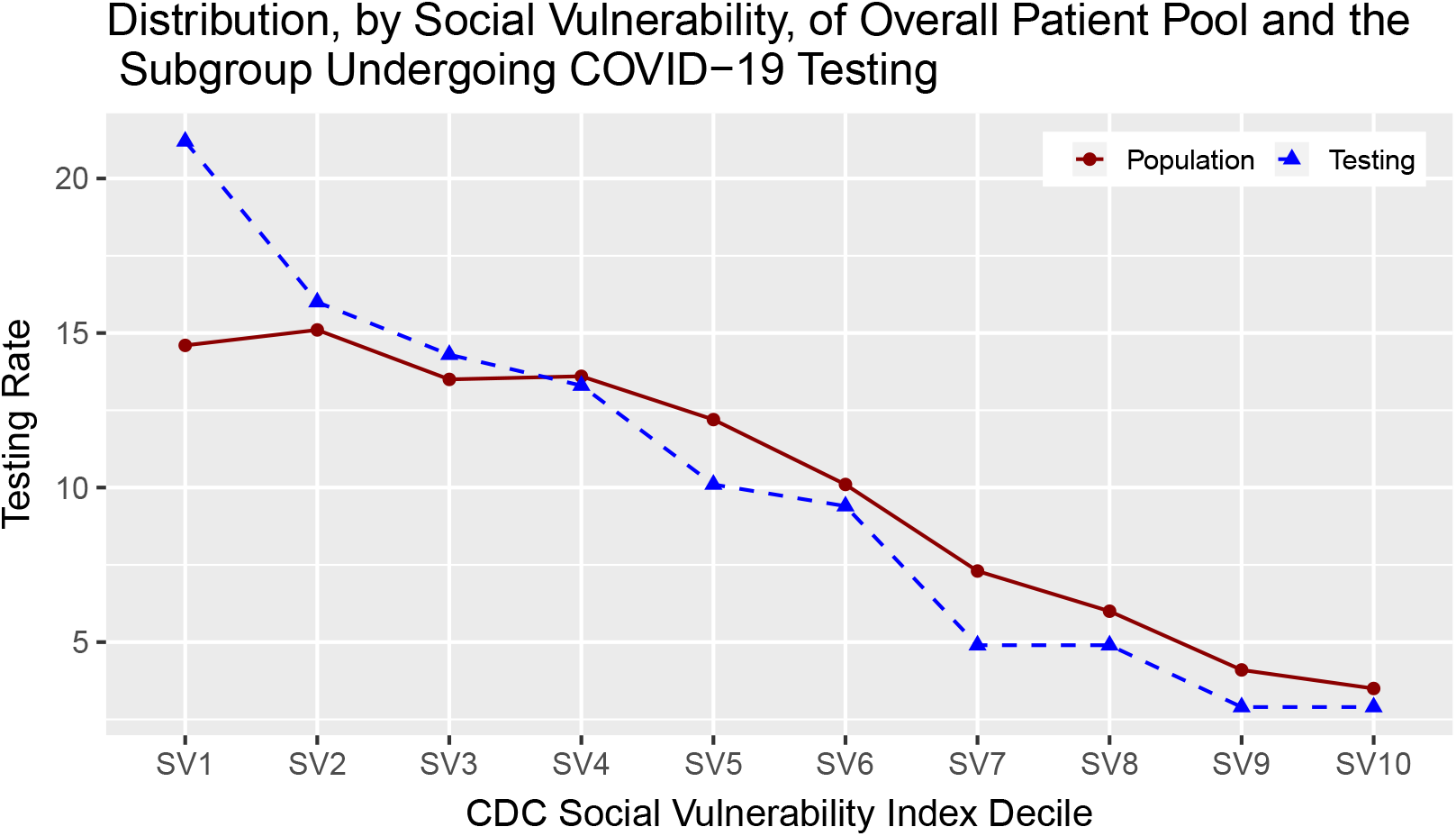
COVID-19 Testing Frequency in Relationship to Social Vulnerability. The distribution of the total population (solid line) and the distribution of the population undergoing COVID-19 testing (dashed line) by SVI decile (SVI 1 = least vulnerable; SVI 10 = most vulnerable).

### COVID-19 Test Positivity

Whereas rates of testing among racial groups were fairly similar to their patient pool proportions, COVID-19 test positivity rates strongly diverged with respect to race and ethnicity. Similar to reports from other studies in the US, in our study population, the Black and Hispanic groups also tested positive at higher rates than their White and Asian counterparts. Hispanics individuals exhibited the highest rate of test positivity (11.76%)—more than double that of Black individuals (5.38%), who had the second highest group rate.

Figure 2 shows COVID-19 testing frequency and test positivity rates in relation to SVI decile. The histogram bars show the total number of tested performed (left-hand-side *y*-axis). The dashed line shows the positivity rate (right-hand-side *y*-axis). We can see from the histogram bars that the number of tests by the least vulnerable populations far exceed the number of tests by the most vulnerable populations. At the same time, the most vulnerable SVI decile tested positive at 4.8 times the rate of the least vulnerable SVI decile. The steadily increasing dashed line shows that the rate of test positivity rises with SVI decile.

**Figure 2:**
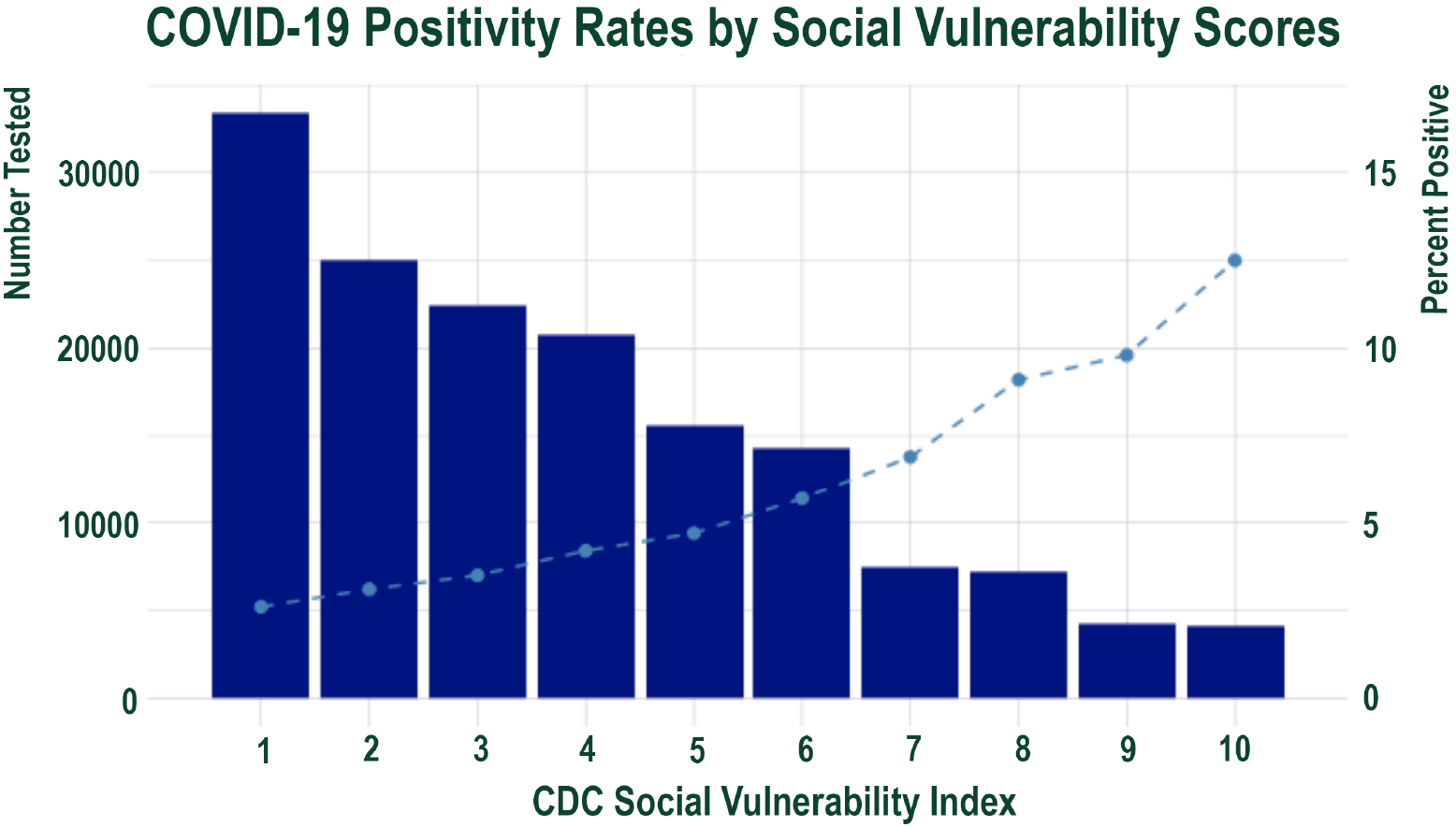
COVID-19 Positivity Rates in Relationship to Social Vulnerability Index. The total number of tests performed is on the left y-axis). The right y-axis shows positivity rate (right y-axis). Individuals with higher social vulnerability test much less frequently than those with lower social vulnerability yet have much higher positivity rates, even when correcting for their lower overall rate of testing.

### COVID-19 Hospitalization Rates

Table 3 lists the rates of hospitalization by race and ethnicity among those testing positive for COVID-19. The hospitalization rate for all age groups and the hospitalization rate for just those aged 60 years and over are displayed separately. To identify COVID-19 hospitalizations, we first identified all hospitalizations. We deemed the hospitalization to be a COVID-19 hospitalization if three conditions were met. First, the hospitalization occurred between February and December 2020. Second, the hospitalization was at least 24 hours. Third, the hospitalization visit included the ICD-10 code U07.1, which indicates that a COVID-19 diagnosis is associated with the hospitalization.

**Table 3:** COVID-19 Hospitalization and Death Rates by Race/Ethnicity and Age.

|  | White |  | Black |  | Hispanic |  | Asian |  |
| --- | --- | --- | --- | --- | --- | --- | --- | --- |
| Hospitalization Rate (All Ages) | 5.36% | (100) | 9.28% | (54) | 6.87% | (187) | 11.46% | (102) |
| Hospitalization Rate (60+ Years) | 10.75% | (70) | 17.45% | (26) | 14.87% | (51) | 26.92% | (70) |
| Death Rate (All Ages) | 0.54% | (10) | 1.24% | (7) | 0.37% | (10) | 1.93% | (17) |
| Death Rate (60+ Years) | 1.53% | (10) | 4.08% | (6) | 1.72% | (6) | 5.77% | (15) |

Absolute number shown in parentheses.

In our data, White patients, who had the lowest test positivity rate (2.5%; Table 2), also had the lowest hospitalization rate (5.36% overall; 10.75% for age 60 and older). In contrast, Asian individuals, despite testing positive at the second lowest rate (3.68%), experienced the highest rate of hospitalization, both overall (11.46%), and in the 60 years and older age group (26.92%). The hospitalization rate for Asian individuals was more than double the corresponding rates for White individuals. Black individuals had the second highest hospitalization rate (9.28% overall and 17.45% for ages 60 and above).

Figure 3 shows hospitalization rates by SVI. While one might expect that hospitalization rates would increase with social vulnerability, a consistent relationship where hospitalization rates generally rose with increasing social vulnerability was observed only for the White group. Hispanic persons experienced largely similar hospitalization rates across all SVI quartiles. For Asian and Black individuals, the relationship between hospitalization rates and SVI was variable. Notably, Asian patients had the highest hospitalization rates overall in all four SVI quartiles.

**Figure 3:**
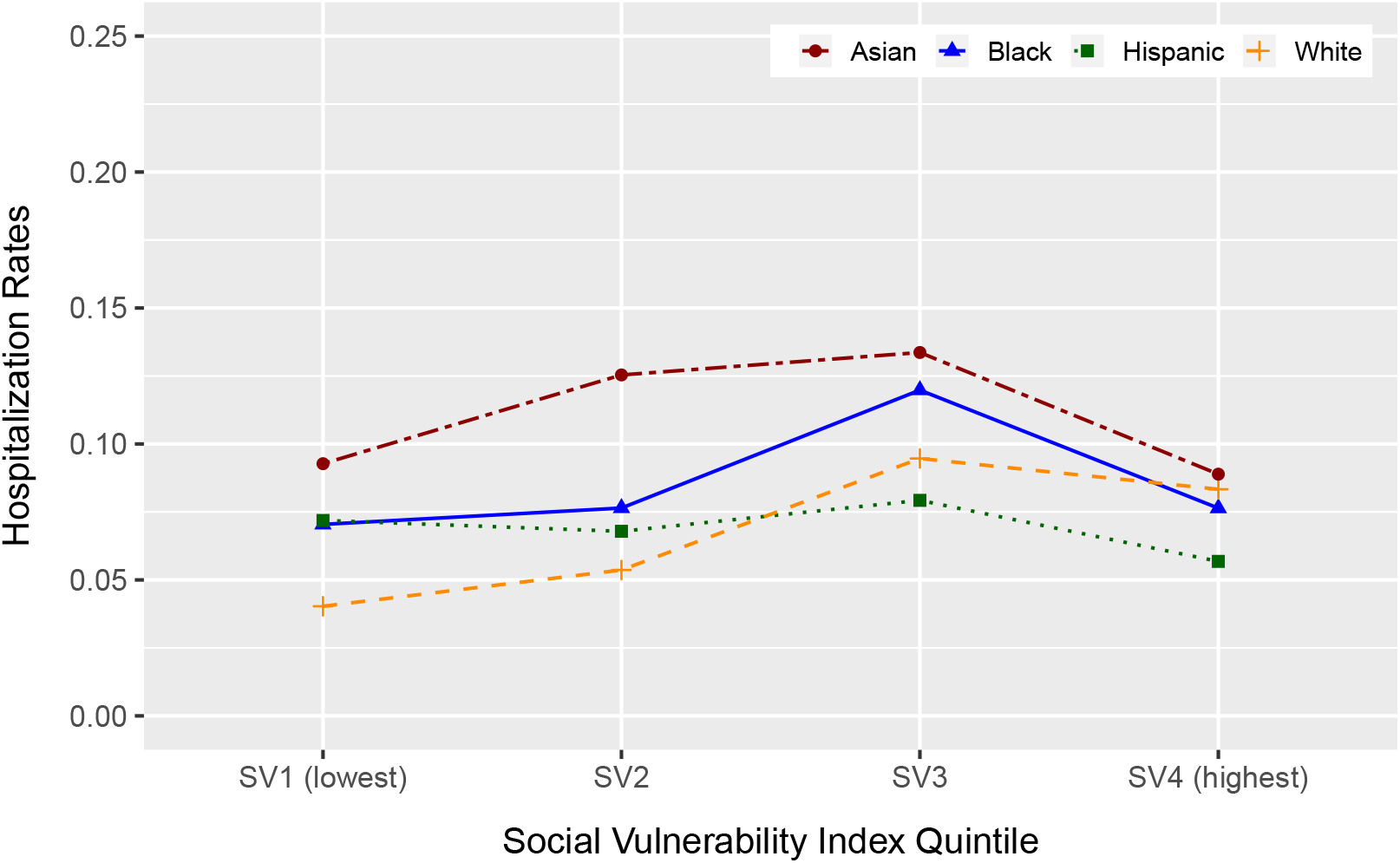
COVID-19 Hospitalization Rates by Race and Social Vulnerability Scores.

### COVID-19 Death Rates

Table 3 also displays COVID-19 death rates by race. Attribution of cause of death to COVID-19 was based on admission diagnosis and associated diagnoses during hospitalization compatible with COVID-19 mortality. Since our data are EHR, further corroboration with death certificate-listed cause of death could not be performed due to masking of patient identity necessitated by confidentiality requirements.

Between the four racial and ethnic groups, Asian individuals exhibited the highest mortality *rate*, both crude and age-adjusted. Asian individuals also had the highest *number* of COVID-19 deaths, both overall and in those aged 60 years and older. The Hispanic group had the lowest crude and second lowest age-adjusted mortality rate, but Figure 4 shows that the Hispanic group also had the lowest mean age at death. For all of the groups, the vast majority of deaths occurred among those age 60 and over, but for White and Asian patients, most of the deaths were among those who were even older (commonly in their 80s and 90s). The average expected years-of-life-lost due to COVID-19 mortality for Hispanic individuals was 25.8 years, far higher, and at least twice as large as for any of the other racial groups.

**Figure 4:**
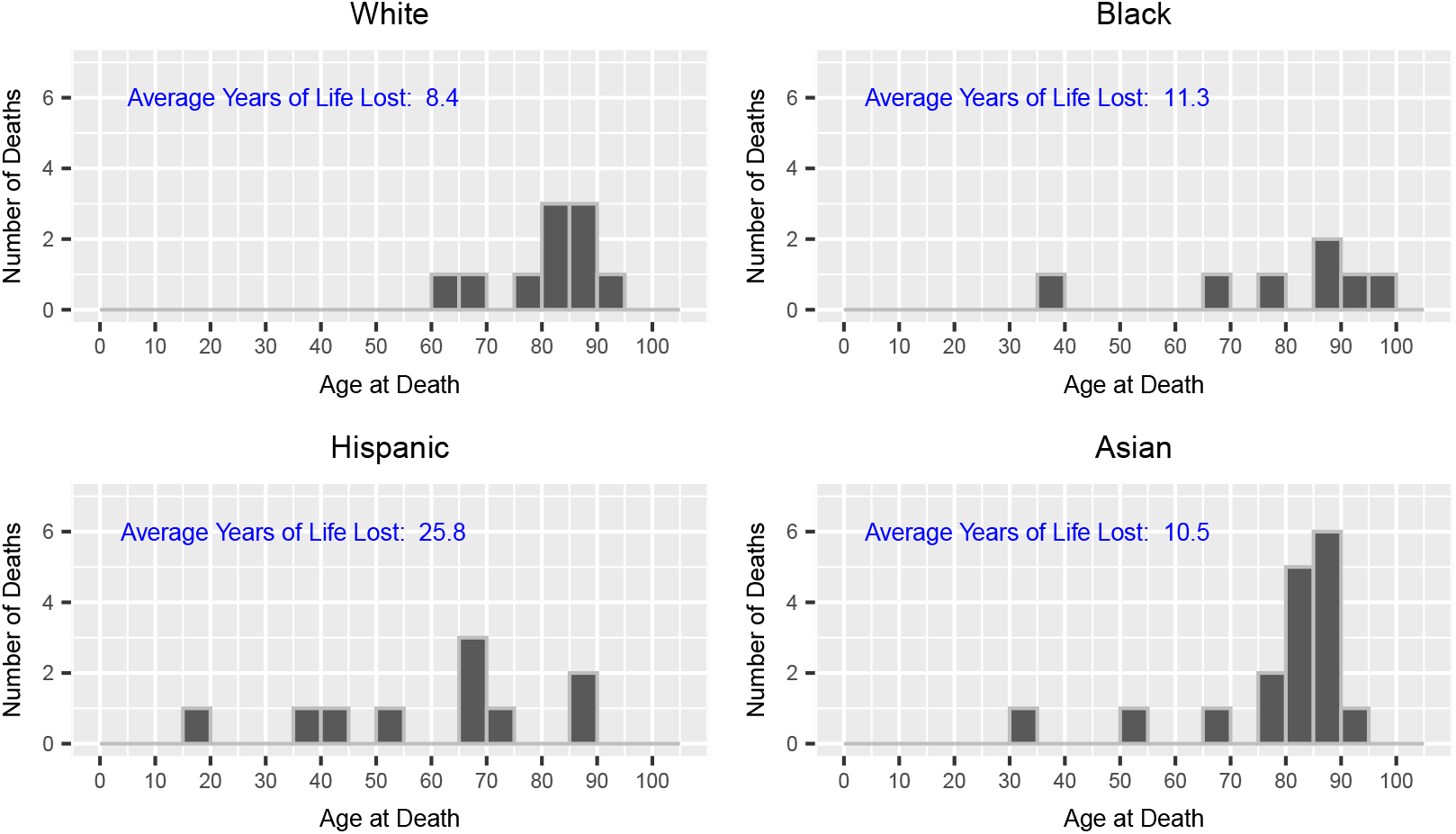
Average Years of Life Lost in Patients Dying of COVID-19, According to Race/Ethnicity.

The way in which we define COVID-19 deaths is conservative and likely misses some COVID-19 deaths. If we adjust the way COVID-19 deaths are assessed so that we allow our definition to include individuals who tested positive with COVID-19 and passed away within 3 months, the number of deaths at least doubles in each of the listed categories in Table 3, and in some cases rises even more. Notably, however, the pattern does not change. With either the conservative definition of COVID-19 death or the more expansive one, Asian individuals exhibit the highest death rates, crude or age-adjusted. As well, the average number of years lost is at least twice as large for the Hispanic group than for any other group. The only noticeable difference is that the more expansive definition of COVID-19 deaths captures more deaths by White individuals. In fact, it captures sufficiently more deaths by White individuals that the White group then becomes the group with the highest *number* of deaths, though the Asian and Black groups still have a higher *rate* of deaths than the White group.

The number of deaths is quite small in our data set, in part because we restricted our time interval to the period of time at the beginning of the pandemic. Given the relatively small numbers of recorded deaths at the beginning of the pandemic, it is impossible to perform tests for statistical significance or to conduct more rigorous analysis of age-adjusted mortality. All the same, the striking patterns observed in this study have not been widely reported previously and deserve corroboration and further study. Although we are unable to rigorously explore an age-adjusted analysis of COVID-19 deaths, we observed that the years of life lost due to COVID-19 mortality was considerably higher for Hispanic individuals than for individuals from any of the other racial and ethnic group.

It is worth noting that the US Hispanic population is considerably younger than other racial and ethnic groups. A report by the Pew Research Center in 2016 found that “… about one-third, or 17.9 million, of the nation’s Hispanic population is younger than 18, and about a quarter, or 14.6 million, of all Hispanics are Millennials (ages 18 to 33 in 2014) … By comparison, half of the black population and 46% of the U.S. Asian population are Millennials or younger. Among whites, the nation’s oldest racial group, only about four-in-ten are Millennials or younger (39%)”[14]. In our study population, as well, the Hispanic age distribution skewed to the left of the age distributions for the other groups. In our data set, the mean age for Hispanic individuals was 34 years, in comparison to Black individuals at 43, White individuals at 52, and Asian individuals at 46. However, even accounting for the younger age distribution of the Hispanic population, the proportion of COVID-19 deaths in young Hispanic individuals is strikingly high. A proper age-adjusted analysis of a larger data set is warranted to explore this observation further.

## Discussion

We examined COVID-19 demographics in a geographic region of the US that facilitated differentiated examination of the relationship between race and socioeconomic status among the four largest racial and ethnic groups. The unique demographics of our study population, which included greater racial and ethnic as well as wider socioeconomic diversity than in most of the US, allowed us to draw some distinctions between the relationship of race and ethnicity and the relationship of social determinants (as encapsulated in the CDC Social Vulnerability Index) on COVID-19 outcomes. Our analysis uncovered multiple instances of discordance between the relationships of race and ethnicity and the relationships of social vulnerability on COVID-19 test positivity rates, morbidity, and mortality.

COVID-19 testing rates were inversely associated with increasing social vulnerability but not with race and ethnicity. While the Black, Hispanic, and Asian groups had hospitalization rates that were higher than the White group, when we separated the data into SVI quartiles, we did not detect a noticeable consistent additional relationship with changes in social vulnerability. Only among White individuals was there noted a relationship between higher social vulnerability and modestly increased risk of hospitalization.

Our study provides yet another data point confirming the well-established finding that Black and Hispanic individuals test positive at higher rates than White individuals in the US [15] as well as in Northern California [16], which includes the Bay Area from which the study population was drawn. Notably, however, we found that Asian individuals in the Bay Area had both the highest hospitalization and the highest mortality rates among the four largest racial and ethnic groups. Importantly, this elevated hospitalization rate was observed to be independent of any relationship with social vulnerability. The observation that US Asian groups have had poorer outcomes during the COVID-19 pandemic has been found in some studies [17, 18, 19], though how they have fared appears to differ depending on context [20], with at least one meta-analysis suggesting paradoxically that Asian individuals with *higher* socioeconomic status were likelier to have poorer outcomes. Clearly, further study is needed.

The heightened impact on the Hispanic group, with the greatest number of potential years of life lost, is also of concern, not widely noted, and in need of further inquiry. Given that this group is significantly younger on average, and that lower rates of morbidity and mortality of COVID-19 are expected for younger individuals, *ceteris paribus*, the high numbers of deaths among Hispanics is troubling. Future research is needed to validate this finding and identify potential underlying factors.

### Study Limitations

Our study conclusions are specific to a vaccine-naive patient population that sought care at UCSF Health in 2020 and may not be generalizable to populations in other settings or at later time points in the pandemic. Furthermore, our study population differed somewhat in racial and ethnic composition from that of the Bay Area population. The reasons for this discrepancy could have reflected the region’s geographic heterogeneity in the distribution of Asian individuals, as well as other factors, including but not limited to variations in patient access to or preference for UCSF Health care facilities, patient willingness to seek medical care, health insurance coverage-dictated limitations, and referral effects. Patients who lived a distance from a UCSF Health facility may have been less likely to seek care at UCSF Health and thus would been less likely to be included in the study population. For example, Santa Clara County, the most populous in the nine-county Bay Area, lies beyond the immediate referral zone of UCSF Health yet has highest percentage of Asian individuals (38.9%) in the region. Furthermore, it is conceivable that any such distance-related access barriers would have been more difficult to overcome for individuals with higher social vulnerability and lower socioeconomic status. At the same time, amongst large, regional health care systems in the San Francisco Bay Area during the study period, UCSF Health cared for a disproportionately greater share of uninsured and underinsured patients, the result of which may have tended to mitigate any potential selection bias against individuals with lower socioeconomic status.

Self-reported residence address data were used for geocoding. We performed manual review and correction of certain errors where possible (e.g., misspelled street names and transposed zip code digits) but excluded records that could not be geocoded (e.g., missing street addresses). These limitations may have introduced some systematic errors, such as underreporting of homeless individuals or miscoding of records in which a mailing address rather than a place of residence had been entered. The magnitude of such errors is unknown, but there is no particular reason to believe that their exclusion would create systematic bias in a particular direction.

Socioeconomic vulnerability was calculated based on census tract data assigned from geocoded address data, a process that, while standard in demographic studies, could have introduced potential attribution error based on differences between group-level measures and individual-specific data. The electronic health record captures only a circumscribed subset of data regarding a given patient’s social determinants, so studies conducted in similar fashion to ours [21] must necessarily rely on group-level rather than individual measurements.

It is possible that the number of COVID-19 positive tests, COVID-19 hospitalizations, and COVID-19-related mortality would be underreported if some patients had undergone negative testing at a UCSF Health site but subsequently underwent further testing, required hospitalization, died at another health care facility, or moved out of the area. To our knowledge, there have been no published studies that would suggest that any such effect would be more likely to affect one racial and ethnic group over another or to preferentially affect individuals of higher or lower social vulnerability.

### Implications and Next Steps

Increased awareness of the finding that Asian individuals are a vulnerable COVID-19 subpopulation should prompt additional study of Asian group-specific causative factors underlying the observed increased rates of COVID-19-related hospitalization and death. Such investigation may not only be beneficial in designing interventions to address health disparities affecting Asian communities, in particular, but could also be potentially critical to identifying and mitigating the root causes of COVID-19-related racial disparities across the entire population. Indeed, the internal diversity of Asian communities and their unique attributes as an aggregated racial group provide valuable opportunities to better understand and hopefully mitigate the generalized problem of COVID-19-related racial disparities in the population as a whole. In particular, studies of socioeconomically diverse Asian populations such as that of the Bay Area may shed fresh insights that could help untangle the interplay of biomedical, social, and other determinants, not only for COVID-19 but potentially other health conditions as well.

Although often lumped together into one racial construct, Asian communities encompass wide diversity in ancestry, culture, religion, language, immigration status, educational attainment, socioeconomic status, and experience with racism (including internal, interpersonal, and structural) [22]. Further study and improved research methodologies, such as data disaggregation and uniform collection of Asian race and Asian subgroup ethnicity data [23, 24, 25], are needed to better understand the factors contributing to the elevated risk for COVID-19 morbidity observed in Bay Area Asian individuals, as a whole. These studies should include more robust collection of individual-specific social determinants of health, further investigation of biomedical risk factors, and identification of relevant factors that may not be captured within current measures of social vulnerability, such as the multiple effects of structural and interpersonal racism [26] that could, as just one example, affect the willingness of Asian individuals to seek or receive equitable and timely health care [27].

Our study period at the beginning of the pandemic limited our ability to rigorously examine COVID-19 deaths. Sadly, many more deaths have occurred since 2020 and many of those deaths have been in minority communities. While a study of a longer time period must contend with the additional and possibly confounding effects from vaccines, mask mandates, and virus variants, it also provides an opportunity to further examine the impact on the Hispanic community. Given their younger population, one would expect their community to better weather this virus that has devasted the elderly population. If this pattern of deaths persists throughout the pandemic, understanding why is a critical component of unraveling the complexity of the COVID-19 racial health disparities.

Finally, while identifying broad patterns of racial disparity in health outcomes and health care at a national level is important and relevant, alleviating structural inequality must occur through specific and intentional measures that are locally and contextually relevant. Accordingly, a comprehensive understanding of COVID-19 impacts on racial and ethnic groups in the US must include study of regions and subpopulations that differ substantially in makeup from the US as a whole, not only to better understand race-specific causation factors, but also to formulate data-driven strategies and policies to ameliorate racial disparities in COVID-19 outcomes. Any such mitigation strategy must consider the US not as a homogeneous entity but as a diverse collection of regional subpopulations, some of which may differ substantially in their manifold demographic attributes, including racial and ethnic composition [28], socioeconomic distribution [28], and health-related characteristics [29], as well as structural aspects such as, for example, housing stock [30, 31], educational opportunities [32], employment characteristics [33], transportation networks [34], access to health care [28], historical evolution of neighborhoods[35], and experience of systemic racism [36].

## Conclusion

The impact of COVID-19 has been felt by all races, albeit in different ways. In the San Francisco Bay Area, Black and Hispanic individuals tested positive at the highest rates, but Asian individuals experienced the highest hospitalization and death *rates*. At the same time, Hispanic individuals suffered the greatest number of potential years of life lost [37] and so experienced disproportionate *impact* from mortality due to COVID-19. To be sure, even while particular effects may vary, no racial minority group has been left unscathed by COVID-19, underscoring the need to not only better understand individual-specific and group-related determinants of health, whether biomedical, social, or otherwise, but also to address racial and racism-related health disparities [38] and thereby positively impact the health of the entire population. Reducing both inter-group disparities as well as the absolute toll of COVID-19 remains an important target for study, intervention, and policy, inasmuch as racial disparities and health inequality impact not just the most vulnerable among us but all strata of our interconnected society.

## Data Availability

Data are available through UCSF with IRB approval

## Acknowledgements

The research was supported in part by unrestricted grants to DGH from All May See (formerly That Man May See) and Research to Prevent Blindness. The authors also wish to thank Ashly Dyke for assistance with the literature review. This work used the Extreme Science and Engineering Discovery Environment (XSEDE), which is supported by National Science Foundation grant number ACI-1548562. Computing resources were granted through the HPC Consortium for access to Pittsburgh Supercomputer Center’s Bridges, Bridges-2, and Bridges-AI.

